# Impairment of T cells’ antiviral and anti-inflammation immunities dominates death from COVID-19

**DOI:** 10.1101/2021.04.26.21256093

**Authors:** Luhao Zhang, Rong Li, Gang Song, Gregory D. Scholes, Zhen-Su She

## Abstract

Clarifying dominant factors determining the immune heterogeneity from non-survivors to survivors is crucial for developing therapeutics and vaccines against COVID-19. The main difficulty is quantitatively analyzing the multi-level clinical data, including viral dynamics, immune response, and tissue damages. Here, we adopt a top-down modelling approach to quantify key functional aspects and their dynamical interplay in the battle between the virus and the immune system, yielding an accurate description of real-time clinical data involving hundreds of patients for the first time. The quantification of antiviral responses demonstrates that, compared to antibodies, T cells play a more dominant role in virus clearance, especially for mild patients (96.5%). Moreover, the anti-inflammatory responses, namely the cytokine inhibition and tissue repair rates, also positively correlate with T cell number and are significantly suppressed in non-survivors. Simulations show that the lack of T cells leads to more significant inflammation, proposing an explanation for the monotonous increase of COVID-19 mortality with age and higher mortality for males. We conclude that T cells play a crucial role in the immunity against COVID-19, which reveals a new direction——improvement of T cell number for advancing current prevention and treatment.

## Introduction

The ongoing COVID-19 pandemic has resulted in over two million deaths worldwide. Therefore, identifying key factors determining the immune heterogeneity from non-survivors to survivors is crucial for the current fight against the pandemic. Past clinical studies have found a series of host factors associated with severe disease or higher mortality via correlation analysis: individual characteristics including older age, male sex, and comorbidities^1,2^; profound lymphopenia, with T cells most significantly affected^3–5^; the elevated level of inflammation markers, like LDH (lactate dehydrogenase) and D-dimer^2,6^; excessive release of pro-inflammatory signalling molecules, like IFN −*γ*, IL-6, etc., known as the cytokine storm which is thought likely to be a major cause of multiorgan failure^4,7^. For immune responses, both SARS-CoV-2 specific T cells and antibodies are observed in COVID-19 patients^6,8^. However, the quantitative role of these factors in antiviral and anti-inflammatory immune responses is unknown, resulting in several unsolved questions about the cause of death and the protective mechanism against virus and inflammation: 1. Are T cells and antibodies helpful or harmful^9,10^, especially in severe patients? What are the relative contributions of T cell and antibody response for antiviral immunity at different stages? 2. What are the main drivers and suppressors for the cytokine storm and multiorgan failure? Most importantly, 3.Are there new directions to overcome the heterogeneity of patients, decay of antibody function, and gene mutation SARS-CoV-2 in efficient therapeutics and vaccine developments?

Beyond correlative analyses, quantitative modelling is a powerful tool to simulate the measured dynamical immune response to reveal the relative importance of different components^11^. However, many recent studies focus on the dynamics of the virus and its interactions with immune responses^12,13^and antiviral drugs^14–23^, without considerations of inflammation which is essential in disease progression. On the other hand, some multiscale simulations ^24–27^ incorporate existing knowledge about the viral dynamics, immune responses (with inflammation) to simulate the clinical outcomes. However, these approaches involve hundreds of model parameters, which have considerable value uncertainties that limit the reliability of predictions and systematic comparisons with clinical data. Therefore, previous studies either include no inflammation or include too many cellular or molecular inflammation components and parameters to clarify the key factors dominating death.

In this work, we adopt a top-down modelling to construct a simple and verifiable model including both antiviral dynamics and inflammation. The model quantifies crucial functional aspects in the virus-immune system battle to overcome the difficulty mentioned above. Here, the battle is classified into three kinds of functional behaviours, namely, the pathogenic function (e.g., virus and inflammation), the protective function (e.g., innate and adaptive immunity), and organ damage. Integrating with the existing clinical and immunological knowledge for COVID-19 patients, we establish a dynamical motif for a small set of crucial functional variables and their interplays. The antiviral inflammation model is used to simulate the systematic progression of COVID-19 patients with 19 parameters that can all be estimated from clinical data. These simulations are validated with real-time clinical data involving hundreds of patients and then evaluate contributions of T cells and antibodies to antiviral immune responses. Subsequently, we quantify the difference of anti-inflammatory immune responses from non-survivors to survivors and clarify their correlations to T cells. Finally, T cells’ dominant role in saving the death of COVID-19 and revelation to new therapeutics and vaccine development are discussed.

### Causal network of the Antiviral-Inflammation Model

The difficulty of previous multiscale simulations^24–26^ due to considerable parameter value uncertainties stems from the fact that, in the bottom-up strategy, the immune response to infectious disease is modelled as a complex network of numerous factors, resulting in the so-called ‘curse of dimensionality’^28^. In contrast, in a recent successful model of a classical complex system, namely, fluid turbulence, one of us has demonstrated that the global motions composed of numerous components typically display a symmetry-breaking which can be quantitatively modelled with finite functional variables, called order functions^29,30^. Here, we use a similar top-down strategy conducted from the human body to the molecular level to quantify immune functional aspects in the virus-immune system battle. At the human-body level, we first decompose the system into three functional classes: the pathogenic function, the protective function, and the organ damage. Then, we specify dominant components at the cellular or molecular level for each functional class, ignoring other components. Therefore, this approach captures, by intuition, some essential features to observed physiological behaviours and has avoided unnecessary complexity, which results in a ‘curse of dimensionality with little clinical meaning.

The model explicitly describes dynamics of five crucial functional quantities that determine COVID-19 progression: virus (*V*) and interleukin 6 (IL-6, *I*) for pathogenic function, effector T cells (*T*_*e*_) and neutralizing antibodies (NAbs, *A*) for protective function, D-dimer (coagulation marker, *S*_d_), and high-sensitivity cardiac troponin I (HSCT, heart injury marker, *S*_h_) as examples for multiorgan damage. Other secondary factors modulate their interactions. Fig.1 shows their interplay following time order from virus dynamics, immune response to inflammation response, with variable names and their mutual interaction represented by Greek letters. Self-replicating virus stimulates innate and adaptive immune cells, which can produce antibodies. Effector T cells and neutralizing antibodies (NAbs) clear virus directly, either by killing infected cells or block the virus from entering into tissue cells; non-neutralizing antibodies (Non-NAbs) combines with innate immune cells (macrophages (Mϕ) and nature killers (NK), etc.) and induce immunoreaction to clear virus indirectly. The activated immune cells secrets cytokines, in which IL-6 has a central role for downstream destructive effect on organs, hence, it increases D-dimer and high-sensitivity cardiac troponin I that characterizes multiorgan failure (MOF). On the other hand, suppressing immune hyperactivation and tissue repair by negative feedback reduces activated immune cells and vessel and heart damage, thus reducing IL-6, D-dimer, and high-sensitivity cardiac troponin I.

**Figure 1.**
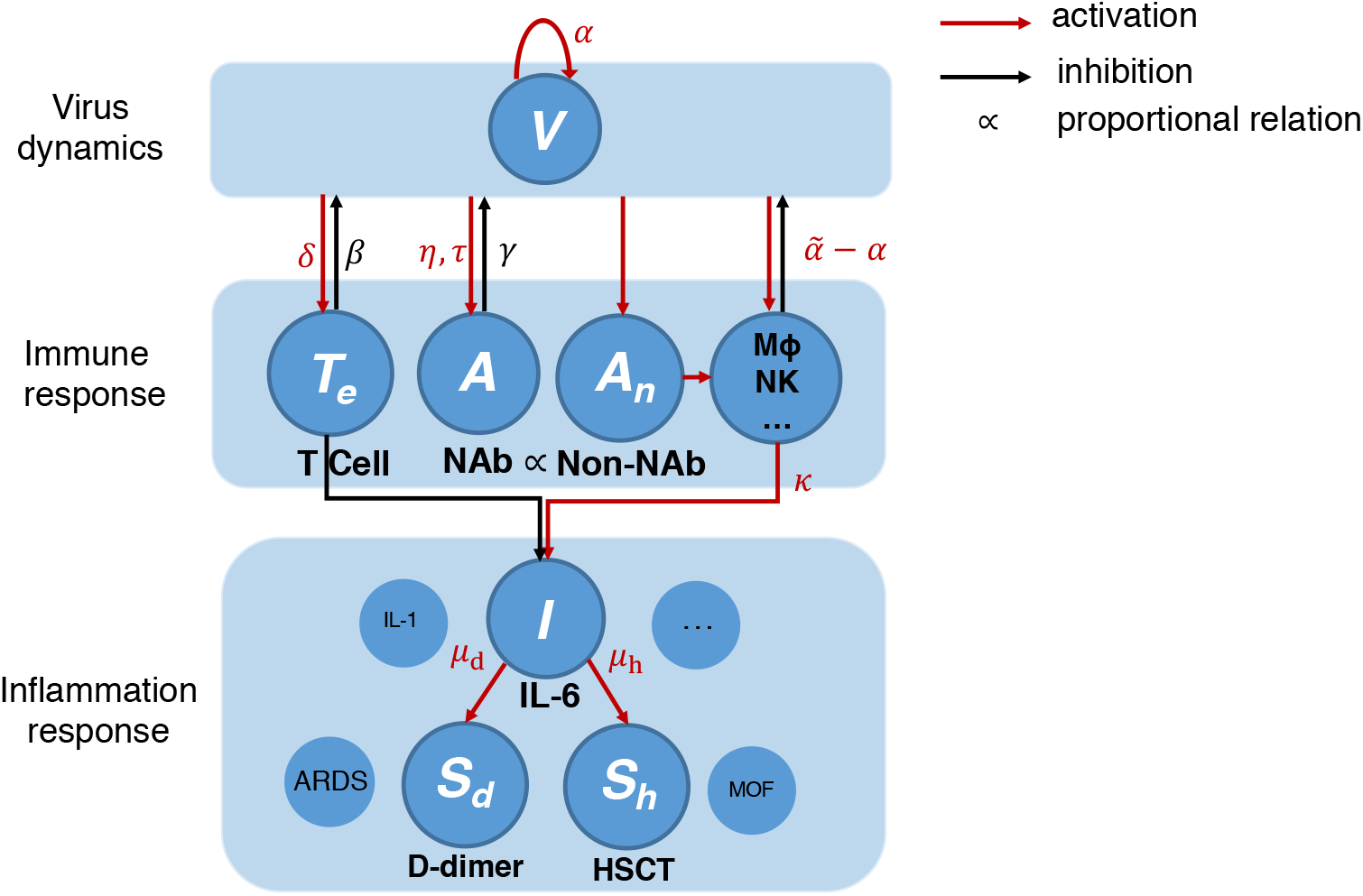
Graphical scheme of COVID-19 antiviral-inflammation model. Key components are highlighted. The red arrows mean activation, and the black arrows represent inhibition. Greek letters mean the activation/inhibition rates or characteristic time associated with each interaction NAb: neutralizing antibody. Non-NAb: non-neutralizing antibody of which the concentration is assumed proportional to NAb. M*ϕ*, NK: macrophage and natural killer cells. IL-6: interleukin 6. IL-1: interleukin 1. D-dimer: coagulation marker. HSCT: High-sensitivity cardiac troponin I, heart injury marker. ARDS: Acute Respiratory Distress Syndrome. MOF: Multi-Organ Failure.

### Mathematical description of the Antiviral-Inflammation Model

Quantitatively, a set of ordinary differential equations, Eq. (1)-(5), is constructed to describe the antiviral immune response and pathogenesis of inflammation at a specific part of the body. The detailed meanings and units of all parameters are provided in SI (Supplementary Information), Table S1.

The antiviral process includes continuous virial replication, dynamical activation of T cells to effector T cells by the virus, dynamical secretion of neutralizing antibodies by B cells, virus clearance by effector T cells and antibodies, and decay of effector T cells and antibodies. Eq. (1)-(3) describes the corresponding dynamical evolutions of concentrations of the free virus (*V*), effector T cells(*T*_*e*_), and neutralizing antibodies(*A*), with concise rate constants and time interval describing global effects of a series of microscopic processes, like target cell infection, antigen identification, antigen presentation, differentiation of T cells, B cell immunoglobulin class switching, etc. 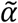 describes an effective viral growth rate, which summates both viral replication rate and viral clearance rate by the innate response. Here, we adopt a constant assumption for 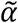, although, in general, it varies in time due to the pathogen lifecycle. This assumption is supported by several studies^13,15,16,21–23^, which showed that, in the early stage, the virus increases exponentially at a single rate, indicating 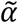 is nearly a constant. While in the later stage, T cell and antibody response significantly^3^, indicating that the innate immunity is unable to control the virus; thus, its variation is negligible. *δ* and *η* represent production rate of effector T cells and antibodies, respectively. Their one-day averaged values are assumed constants compared to the significant change of viral load across several magnitudes. A delay of antibody response that antibodies rise on the 5-15^th^ day after onset^31–33^ is observed, later than the viral load peak. On the contrary, the maximum variation of the T cell concentration is observed to be close to the viral load peak location^3^, which reveals its delay time may be shorter. Therefore, Eq. (3) for antibody response includes a delay, while Eq. (2) for T cell response does not. Besides, in common sense, there is no virus-specific effective T cell or antibody before infection^34^; therefore, *T*_e_(0) and *A*(0) should be 0.

Eq. (4) describes the release and suppression of IL-6. Its production in COVID-19 has two stages; namely, the early inflammation begins after the infection, and the latter inflammation dynamics stimulated by the antibody response. Our simulation focuses on the second stage dynamics for its intimate correlation with severe disease, as shown by IL-6 explosion of non-survivors at 13th day after onset^35^. Therefore, only one production term *κA*(*t*) is included, followed by a negative feedback term with the normal value before infection^3^ represent by *I*_0_. Eq. (5) describes the release of IL-6 causes damage to organs, like thrombus formation and heart injury marked by D-dimer and high-sensitivity cardiac troponin I. The construction is based on the common view that cytokine storm is very likely to cause multiorgan failure^7^. The organ damage marker levels are reduced when blood vessels and the heart are repaired through negative feedback, shown in Eq. (5).

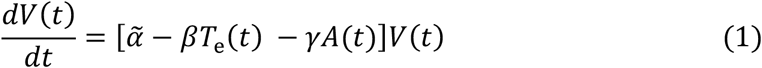

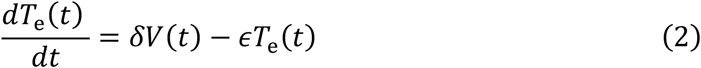

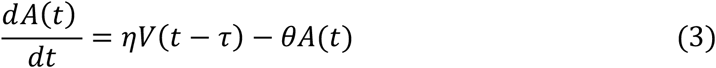

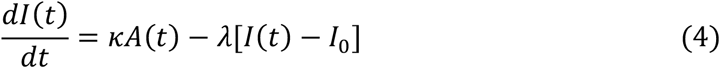

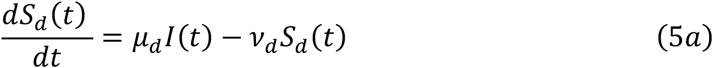

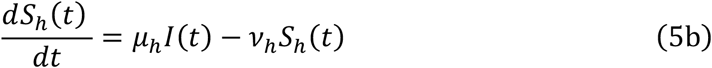

Simulations of Eq. (1)-(5) are compared to real-time data with 457 patients involved. 10 are individuals, and the other 447 patients form mild, severe, survivor, and non-survivor groups. For individuals, we fit each patient’s data directly by Eq. (1)-(5). While for groups, we fit the median values of each variable by Eq. (1)-(5) for we have shown that the group dynamics of patients satisfy a similar set of equations with ensemble-averaged parameters (see SI). For detailed data sources and integration of data from different sources, see Methods and SI. We perform the least-square fit of data using the *fmincon* function of MATLAB with the implemented interior-point optimization algorithm and perform numerical simulation using the delayed differential equations (DDE). The objective function for Eq. (1)-(3), Eq. (4)-(5) are shown by Eq. (10)-(12) in Methods. The time axis for simulation is the number of days after symptom onset, with the starting day being 0 or several days earlier. For a detailed description of the fitting procedure, see Methods and SI.

## Results

### Viral dynamics and contributions of T cells and antibodies to antiviral responses

Virus, effector T cell, and antibody dynamics are simulated to compare with real-time data from 10 individuals^31,36^ and median values of mild and severe (critical) groups, survivors, and non-survivors^35,37,38^. The concentrations of virus (*V*) and antibodies (*A*) are assumed proportional to viral load measurement from the respiratory tract and optical density or titer of Anti-RBD IgG/Anti-S1 IgG/Anti-NP IgG. For T cell data, due to the fact that it’s difficult to obtain systematic data (data that have continuity along time, broad severity spectrum, and a large number of patients) of effector T cells, we use CD3+ T data in serum from Zhang et al.^3^ We classify two categories of T cells: the effector T cells that function in the organism and the T cells remaining in serum. Zhang et al.^3^, argued that the decreased T cells remaining in serum move from serum to organs. Accordingly, we assume that the simulated effector T cell concentration in organs (*T*_e_) is proportional to the reduction of T cell in serum (*T*_serum_(*t*)) from its initial value (*T*_0_): *T*_e_(*t*) ∝ *T*_0_ – *T*_serum_(*t*). In Figure 2a-c, *T*_0_ and *T*_serum_ come from Ref^36^, while in 2d they come from Ref.^3^. Besides, in Figure 2a-c, the T cell count is mapped from the original lymphocyte count is scaled by 0.589 (Methods) for. Besides, when comparing simulation to antibody data, an instrumental baseline value *B*_0_ (usually observed in the experiment)^31,34,37^ is added to *A*(t). For the integration of data sources for individuals and groups, see Methods.

**Figure 2.**
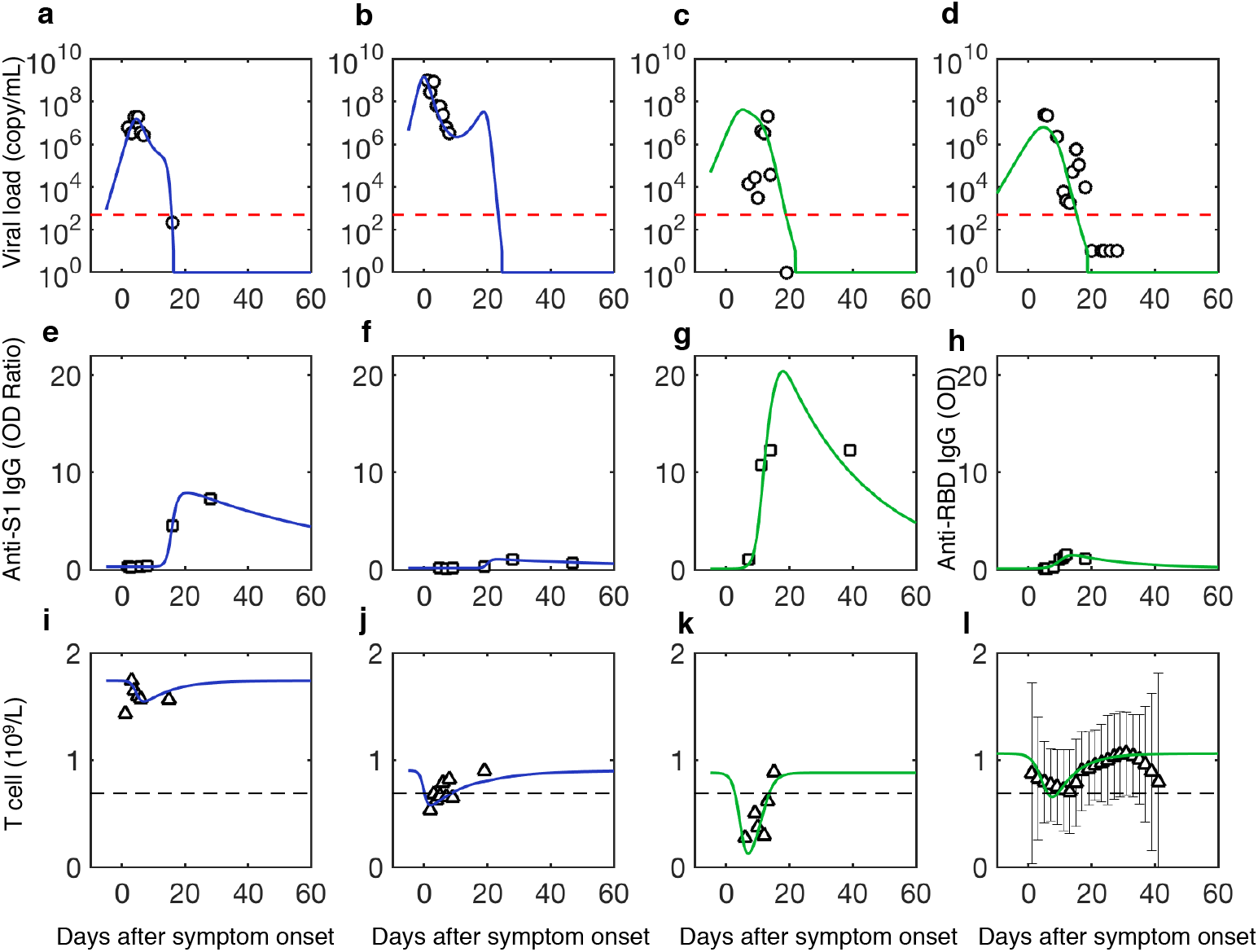
Viral dynamics, adaptive immune response, and contributions of T cells to antiviral responses. a, e, i are fits of the first patient (patient ID labelled as P1); b, f, j are fits of the second patient (patient ID labelled as P3); c, g, k are fits of the third patient (patient ID labelled as P5)^36^; d, h, l are fits of the fourth patient (patient ID labelled as 902). Because the fourth patient lacks T cell data, we use T cell data from severe group^3^ as the T cell value for the fourth patient. Mild patients are in blue and severe patients are in green. Red dotted lines are the limit of detection^31^. Black dotted lines are normal ranges^3^. Viral load is from the nasopharyngeal swab.

As shown in Figure 2 and Supplementary Figure 1, simulations show agreement to data for all cases; for parameters, parameter uncertainties, and the goodness of fit, see SI. Though the fit is done separately for each individual/group with different initial guesses due to the limited number of data points and large fluctuations, the parameters for different patients and groups are within the same order of magnitude (for example, 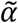varies from 0.41 to 1.86). The goodness of fit, defined as 1 - root mean square/maximum of the simulation of the variable. The mean of the goodness of fit among all variables and patients is 91.8%± 6.6%. Table S8-9 for parameters and goodness of fits of each case. This stability implies that the fitting approach is credible.

Clarifying deterministic factors controlling viral load peak benefits early antiviral treatment, vaccination, and epidemiological control^39^. To understand the main factors that determine viral peak, by asymptotic analysis (Methods), we get an analytical solution that predicts the peak is determined from virus inhibition by T cells: peak value is the ratio between square of virus replication rate, 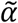and two times the multiplication of T cell activation and virus clearance rate, *δ, β*. This gives 10^7.14^ copy/mL for the viral peak of patient P1 in Figure 2 consistent with the simulated value 10^7.18^ copy/mL. Table S3 in SI summarizes the analytical viral peak values compared to the corresponding simulation, which have >90% overlap with each other for all cases. Data agreement gives that for patients who survive, on average, 95% of viruses are cleared per day with 10^9^/L T cells from blood engaged, revealing strong efficiency of T cells’ virus clearance. In conclusion, the consistency between simulation and data clarifies the antiviral dynamics for various severities in which adaptive response plays a significant role---first, effector T cells are activated, kill infected cells, and induce viral peak; then neutralizing antibodies are secreted, finally, clear the virus.

To clarify the roles of T cells and antibodies in the antiviral process, we define the amount of virus cleared by T cells, *N*_T_ and antibodies, *N*_A_, are: 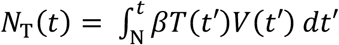 and 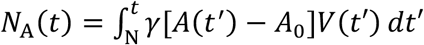. Then, the contribution by T cells in adaptive response for clearing virus, *F*_T_, and the contribution of antibodies, *F*_A_, are: *F*_T_ = ∫ *N*_T_(*t*)*dt*/[∫ [*N*_T_(*t*) + *N*_A_(*t*)]*dt*], *F*_A_ = 1 – *F*_T_.

For patients of different severities, we compare the quantitative contributions of T cells and neutralizing antibodies for virus clearance, as displayed in Figure 3. It shows T-cell immunity dominates the total virus clearance for all patients (88.8%) but significantly decreases from mild to severe patients, consistent with previously reported less CD4+, CD8+ response in severe patients compared to mild patients^8^. Instead, the antibodies’ contributions are 3.3% (mild), 19.4% (severe or critical) and 28.9% (non-survivors), respectively. Our simulation finds that the antibody preparation time before secretion is overall smaller in severe cases (9.42, 5.40-12.79 day) than in mild cases (14.68, 8.45-20.13 day), revealing antibodies in severe patients secrete earlier and cleared more virus. In conclusion, we demonstrate in COVID-19 that T cells have a dominant role in the virus clearance relative to antibodies, especially for mild patients.

**Figure 3.**
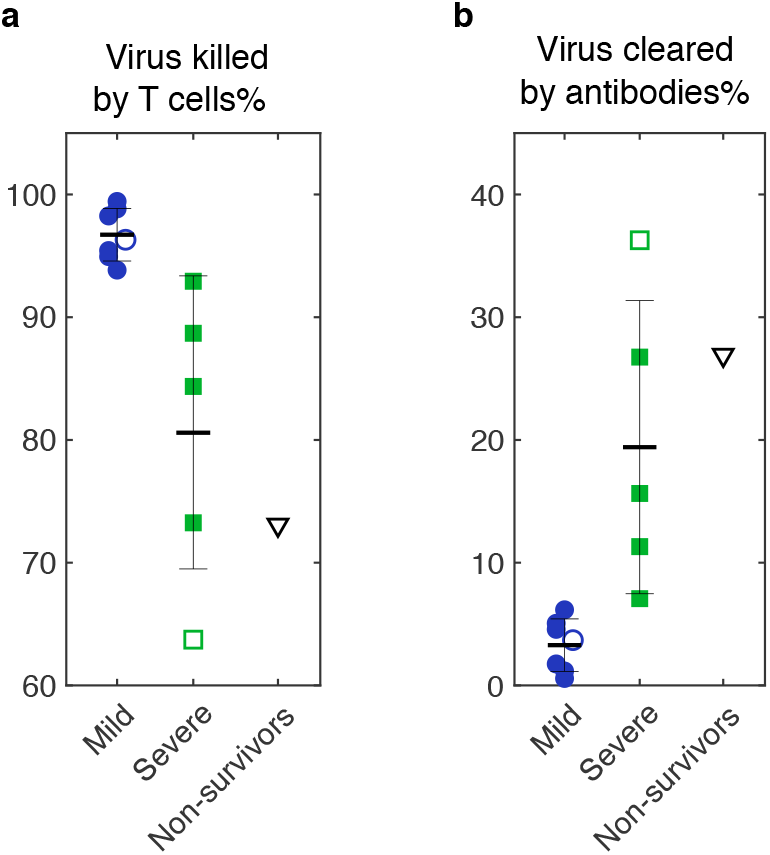
An overall statistic of the fraction of virus killed by T cells (a) and antibodies (b) for all cases. Solid markers are individual data, and hollow markers are group data. Error bars represent standard errors.

### Inflammation dynamics associated with death

To clarify the main driver for the cytokine storm and organ damage of critical illness, we compare simulations of Eq. (1)-(5) with real-time, median data of survivors and non-survivors (Figure 4). The concentration of non-neutralizing antibodies is assumed proportional to anti-RBD IgG optical density. For data source and parameter estimation, see Methods and SI. The agreement between simulations and data of IL-6, D-dimer, and high sensitive cardiac troponin I. The agreement between experiment and group demonstrates the validity of Eq. (1)-(5) and allows us to investigate the critical difference between survivors and non-survivors.

**Figure 4.**
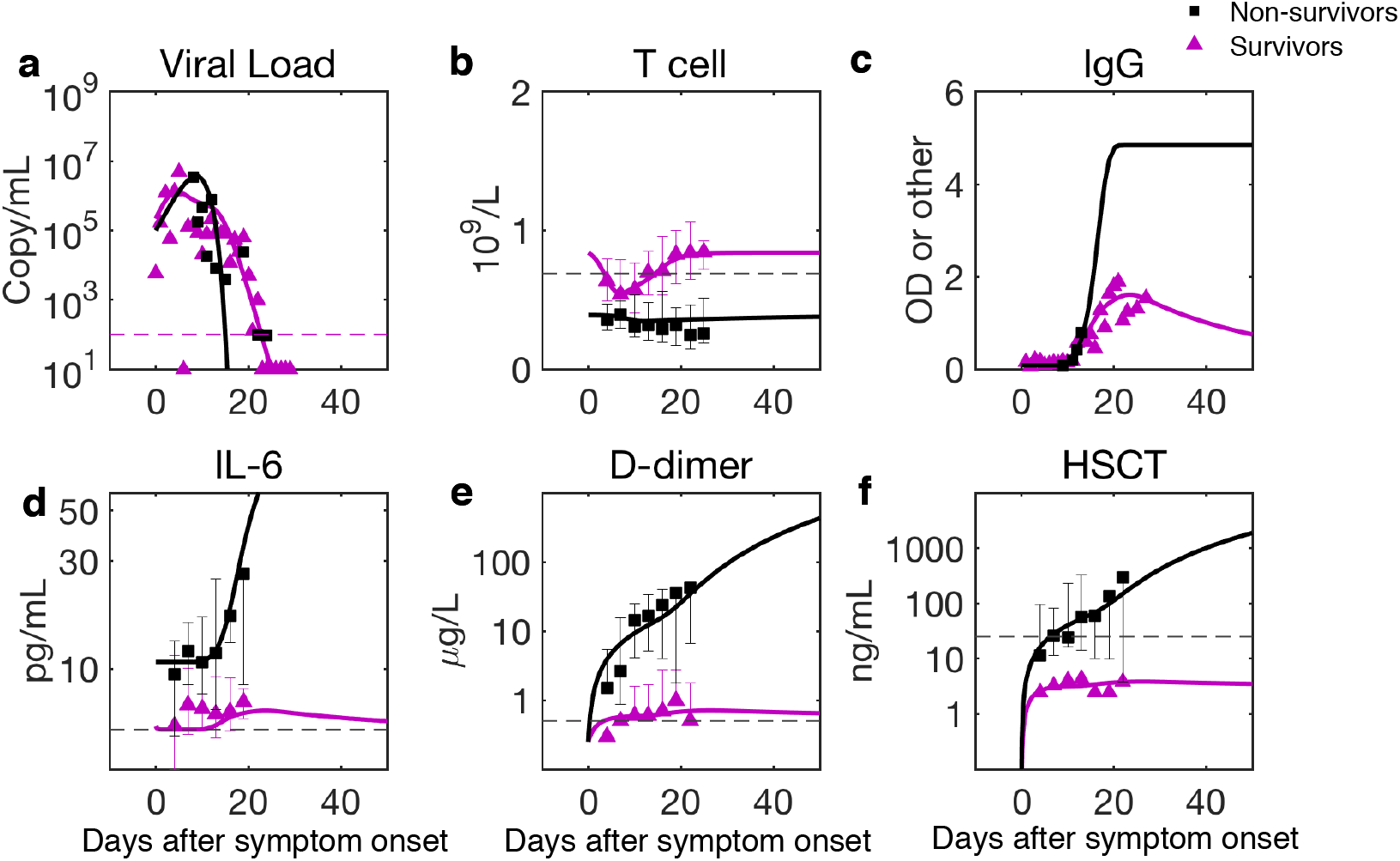
Comparison of predictions to clinical data of survivors and non-survivors. The data are the median of the group with error bars^35^. For parameter estimation, see Methods and SI. The saturation value of Anti-RBD IgG for non-survivors is estimated by assuming its ratio to the maximum of survivors’(18 non-critical and three critical) data is close to the ratio of maximums of neutralizing antibodies between critical and non-critical patients^40^.

IL-6 formation rates are assumed to be the same for both groups. The striking feature of non-survivors compared to survivors is the continuous production of IL-6 and organ damage, revealed by zero inhibition rates for all three markers (Figure 5b), while the difference of formation rates of organ damage markers is not remarkable. Our finding reveals that the crucial aspect of death from COVID-19 is the lack of negative feedback for anti-inflammatory cytokine inhibition and tissue repair. Besides, the initial IL-6 value for survivors is lower than that of non-survivors, which may result from the first stage of IL-6 dynamics where non-survivors lack control ability.

**Figure 5.**
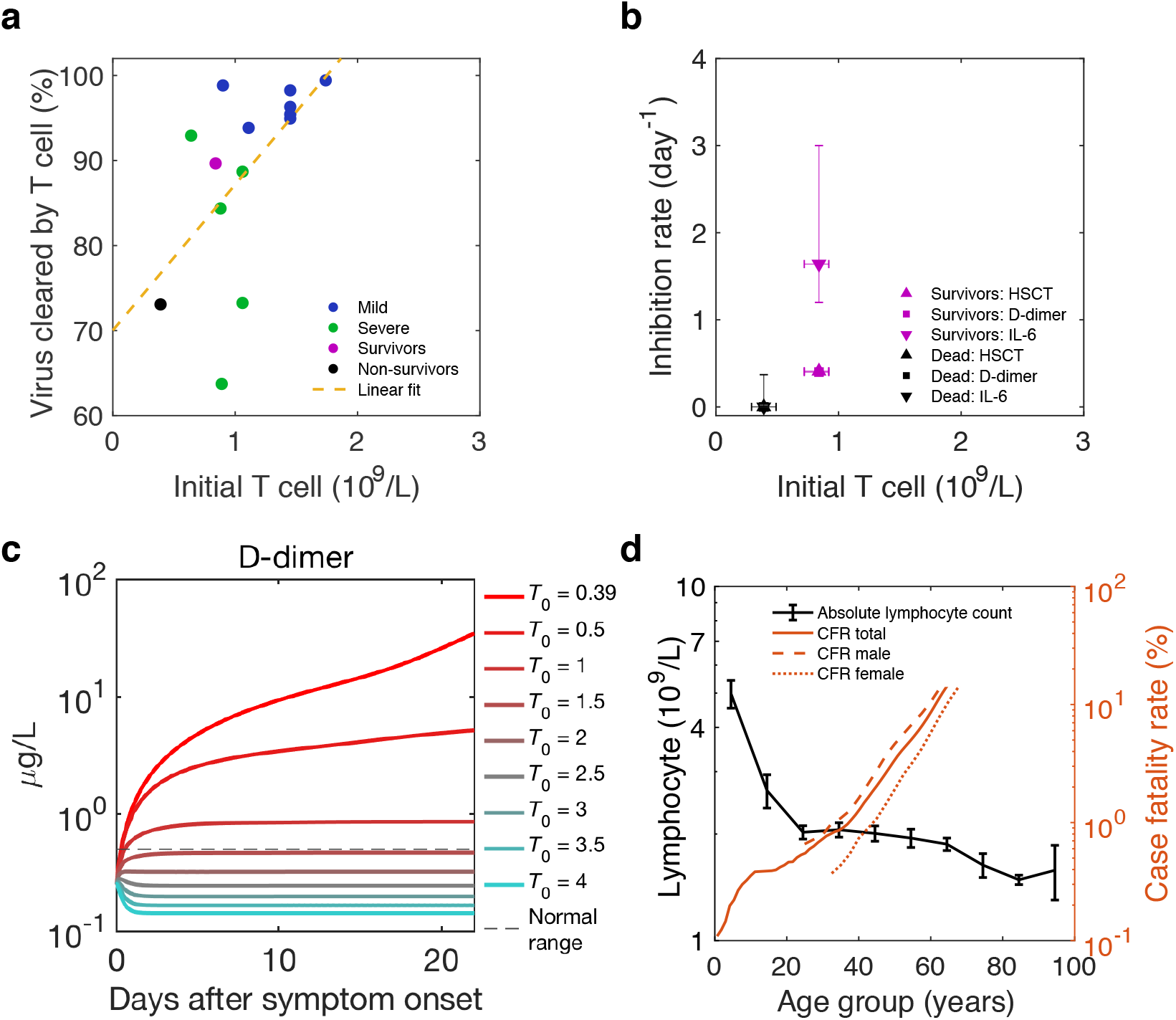
Initial T cell concentration as the background immunity of individuals against SARS-CoV-2 and reduces mortality. a and b: T cell’s antiviral contribution, IL-6, D-dimer, and high sensitive cardiac troponin (HSCT) inhibition rates are positively correlated with initial T cell concentration. R square of the linear fit in Figure 5a is 0.33. c: D-dimer dynamics of non-survivors with an increase of initial T cell concentration (*T*_0_) reduces organ damage at the late stage. The Red dashed line is the normal upper limit of the D-dimer, and the grey dashed line is the median value of the non-survivor group from reference^35^. For parameters of the simulation, see Methods. d. Lymphocyte count decreases with age and mortality (case fatality rate, CFR) increases with age. Male (dashed line) have higher mortality than female (solid line).

### Initial T cells as background immunity that reduce mortality

Our model reveals that virus clearance by T cells, cytokine inhibition, and tissue repair are three essential protective functions in COVID-19 that determine disease severity. To seek what determines these protective functions, T cells’ contribution of total virus clearance, cytokine inhibition rate, and tissue repair rates are plotted with initial T cell concentration before infection (equal to T cell baseline, *T*_0_) in Figure 5a,b, which shows a positive correlation (Figure 5b is based on statistics of 137 non-survivors and 54 survivors). It shows the importance of sufficient initial T cells for comprehensive protection, which comes from an adequate number of effective CD8+ effector T cells that kill infected cells, sufficient regulatory T cells, and other subsets that suppress the immune response and promote tissue repair^41^ to reduce over inflammation in non-survivors.

A great public concern is how an individual patient’s background ‘immune health’ landscape (simplified as background immunity) shapes responses to SARS-CoV-2 infection^9^ and controls the disease’s severity. Because of the determining relation of initial T cell concentration for protective functions, T cells’ static reserve before infection and dominance in population compared to other cells against the virus, we propose concentration of initial T cells is a crucial characterization for the background immunity against SARS-CoV-2. To verify this hypothesis, we conduct disease progression of patients with different initial T cell concentrations. According to Figure 5a,b, by assuming a linear decreasing of T cells’ virus clearing rate, IL-6 and D-dimer inhibition rates with decreasing initial T cell concentration, Fig. 4c shows coagulation becomes more and more significant, which means lack of initial T cells exacerbates disease severity and increase mortality risk.

Therefore, we conclude that the T cells’ impaired antiviral and anti-inflammation functions are the main immune origin of death from COVID-19: the extremely low level of initial T cells in non-survivors results in weak antiviral, cytokine inhibition and tissue repair abilities as well as low tissue repair function; then it calls the elevated antibodies for compensation; as a result, the concomitant large amount of non-neutralizing antibodies amplifies the cytokine storm, leading to continued damage. Following this casual chain, according to the decrease of lymphocytes (hence decrease of T cells assuming T cell count proportional to lymphocyte count) with older age (Figure 5d), we predict straightforwardly older patients must have higher mortality than younger patients. Also, male patients should have higher mortality than female patients for their lower CD4+ T cells^37^. Our prediction proposes an explanation for the continuous increase of COVID-19 mortality with age and higher mortality for males^42^, shown in Fig. 4d. More clinical data are needed to test the validity of the explanation.

## Discussion

In conclusion, we have quantified the adaptive-immune-response heterogeneity from non-survivors to survivors of COVID-19, using a dynamical motif with 19 measurable parameters beyond the overcomplication of the previous multiscale model^26^. For the first time, this model provides an accurate description of real-time clinical data involving hundreds of patients, which then reliably clarifies T cells’ dominant roles in the antiviral and anti-inflammatory immune responses. Furthermore, beyond the previous correlation analysis for T cell scarcity and disease severity^8,9^, this work reveals the causal relation between death from COVID-19 and impaired T cell immunity, provides an explanation for the high mortality of older men.

According to our discussion (Figure 4a, b) of mild and severe patients, survivors and non-survivors, a better vaccine or treatment require better protective functions, i.e., higher virus clearance rate, cytokine inhibition rate, and tissue repair rate, either before or during the infection. Our simulation results in Figure 4c indicate that increasing initial T cell concentration (*T*_0_) can yield more active T cells and, thus, better protective functions and outcomes (e.g., lower D-dimer level at a later stage). Therefore, prevention and treatment approaches that improve (active) T cell number before or during the infection are expected to give better efficacy.

The currently tested drugs target various pathogenesis levels, from antiviral to anti-inflammatory drugs and antithrombotic agents^26^, etc. However, there is no proven effective therapeutics for COVID-19. One crucial challenge is the lack of broad applicability of these drugs to heterogeneous patients with various comorbidities, disease severities, and complications^43^. Our study reveals a new direction will be increasing T cell number and functions by both drugs and health care activities, which may benefit virus clearance, cytokine inhibition, and tissue repair simultaneously. Firstly, recent studies reported the curing effect of drugs to COVID-19 patients by increasing T cell number, e.g., recombinant human granulocyte colony-stimulating factor^44^ JinHuaQingGanKeLi^45^. Therefore, we encourage further studies and applications in this direction. Second, for the recovery of COVID-19 patients and healthy people’s prevention, improving background immunity associated with T cells is more important and promising. Therefore, we strongly suggest studying the curing and immunity improvement effects of health care activities, such as mediation^46,47^, Tai Chi^48^ and BaDuanJin^49^, for previous studies have found they help to increase CD3+ T cell and CD4+ T cell concentration and apply to a wide range of age, including older adults.

On the other hand, in the current development of the vaccine, neutralizing antibody immunity plays a crucial role. Unfortunately, the single-strand RNA structure makes SARS-CoV-2 easy to mutate, and several lineages have been discovered^50^. These mutations pose a challenge for the long-term effectiveness of antibody immunity, for it is on the molecular level targeting specific epitopes of the virus. By contrast, memory T cells show strong cross-reactivity and persistence^34^, and active T cells protect bodies in several aspects, including antivirus, suppress immune hyperactivation, and promote tissue repair. Therefore, stimulation of T cell response by the vaccine is worth more exploration, and we suggest advancing the current combination adjuvant strategy^51^ that elicits potent CD8 and CD4 T cell responses.

For clinical application, to maximize the curing effect for severe patients, we suggest adopting multistage, synthetic protocols incorporating the above therapies. In this case, our model provides a strong tool to evaluate the effectiveness of treatments to identify individual optimal protocols. The reason is that all parameters can be determined from clinical data and quickly predict individual patients’ trajectories, which may also advance the early prediction algorithm of current artificial intelligence softwares^52,53^.

To separate the critical elements from irrelevant details, we here have made some assumptions, which should be evaluated in further clinical studies, although they would not affect the basic conclusions of the study. For instance, the virus-clearance rate of innate immunity is thought to be a constant with negligible variation and is small compared to rates of adaptive response, which should be verified by further time-dependent measurement of the course of the innate response. The second assumption that needs more measurement to test is that, for survivors, the concentration of effector CD8+ T cells at the infected part is proportional to the reduction of CD3+ T cells in peripheral blood; whether it is strictly obeyed at the most time or not might be intriguing to further investigation^4^. The third questionable assumption is that the temporal profile of non-neutralizing antibodies is proportional to neutralizing antibodies. Further measurement needs to test whether they are secreting at the same pace or not.

## Methods

### Extraction of data from published literature

A software tool WebPlotDigitizer (https://automeris.io/WebPlotDigitizer), was used to extract data from fig.2 in ref^3^, fig.1, and fig.3 in ref^36^, fig.2 in ref^2^, fig.1 and fig.3 in ref^37^ and fig.3 in ref^38^. All extracted data were made available to readers in our GitHub shared folder: https://github.com/luhaozhang/covid19

### Data source and integration of data from different sources

There are in total 10 individuals and 4 groups in this work. 4 patients are from Isabella Eckerle’s cohort^36^ and 6 patients from Kelvin To’s cohort^54^, and the severity classification follows the assignments in previous publications. One patient from Kelvin To’s cohort who has not yet been identified as being critically ill in the original paper is classified as severe because the probability of being critically ill is low (1/6). In all, there are 6 mild and 4 severe (including critical) patients. The four groups are mild, severe (including critical), survivors and non-survivors.

The viral load data are from the oropharyngeal swab/posterior oropharyngeal sample /endotracheal aspirates sample measured during the 0th to 30th day after onset. T cell, Lymphocyte, antibody, IL-6, D-dimer, and HSCT are measured from serum sample during the 0th to 30th day after onset. Lymphocyte data were multiplied by 0.589 to estimate T cell concentration (0.589 is the ratio between medians of normal ranges of T cells^3^ and lymphocytes^36^. See Table S2 for how the virus, T cell, and antibody data for all individuals and groups are integrated from various data sources. The IL-6, D-dimer, and HSCT data for survivors and non-survivors are all from BinCao’s cohort^55^.

### Least square fit of virus, immune response and inflammation data

For parameters of simulations in Figure 2 and Figure 4, we adopt a best-fit approach to find the parameters which minimize the given objective function: the mean of residual sum of squares (RSM) between data points and the corresponding model simulations as used similarly in influenza model^56^. For virus-T cell-antibody dynamics, the objective function is:

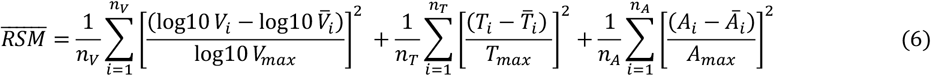

*V*_*i*_, *T*_*i*_, *A*_*i*_ represent values of viral load data, T cell count data and antibody data, respectively. 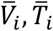 and *Ā*_*i*_ represents the corresponding simulated viral, T cell and antibody value given by our model, respectively. *V*_*max*_, *T*_*max*_ and *A*_*max*_ represent the maximum value among viral load data, T cell count data and antibody data, respectively. *n*_*V*_, *n*_*T*_, *n*_*A*_ are the total number of viral load, T cell count and antibody data points. For the objective function of inflammation response, the mean of RSM (Eq. (7)) was used in linear scale for survivors and log scale (Eq. (8)) for non-survivors. *I*_*i*_,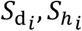 represent values of IL-6 data, D-dimer data and HSCT (High-sensitivity cardiac troponin I) data, respectively and *Ī*_*i*_,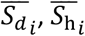 are the corresponding simulated IL-6, D-dimer and HSCT value by our model. *I*_*max*_, 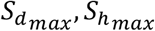 represent the maximum value among IL-6, D-dimer and HSCT value.

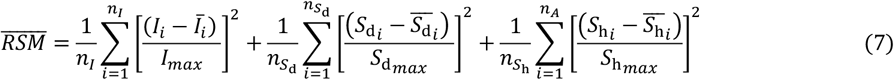

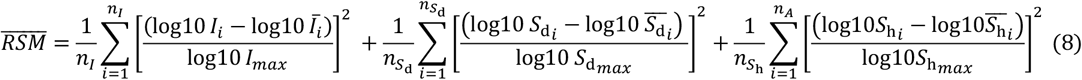

For simulation of viral load dynamics, when *V*<10 copy/mL, it is thought to be cleared thoroughly at one time without further evolution and is set to be 1 copy/mL. The fit that minimizes the objective function with largely fluctuating data points eliminated is called the best fit, which gives results in Figure 2 and Figure 4. Table S5 lists the data points used for performing the best fit of each case. The principles that we use to eliminate largely fluctuating data points are: For virus, data points that represent negative results of the virus are abandoned unless they were important indicators of the ending of viral activity; the data points associated with the second viral load peak in mild and severe group are abandoned because the phenomenon is not observed as the common feature of individual patients. The decay of CD3+ T cells of 902 and 910 patients after 30 days is not used for fit because of the large 95% CI of the data.

*fmincon* function of MATLAB (MathWorks, version 2012 and higher) with the implemented interior-point optimization algorithm is used to perform the fits. It requires constraints for the parameters to be optimized and an initial guess. An empirical fitting is performed for each patient and group to identify initial guess and parameters’ constraints for optimization, shown in Table S10 and Table S11. Patients with the same type of data and same severity category are set to have similar parameter ranges for optimization. A random initial guess is not suitable here because the fit is sensitive to the initial space, probably because of the limited number of data points with relatively large fluctuation, especially for viral load. For fits of survivors and non-survivors, we first fit virus, T cell, and antibody data, then fix relevant parameters and perform fit of IL-6, D-dimer, and HSCT data.

Estimation of the uncertainty of parameters is carried out after the best fit for each case. For parameters related to viral dynamics,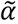, *V*_0_, *β, γ, δ, η, τ*, and inflammation-related rates, *κ, λ, μ*_h_, *v*_h_, *μ*_d_, *v*_d_, we use a similar approach to estimate parameter uncertainty as that in Marchingo, J. M. et al. Science. 346, 1123–1127 (2014). They use an artificial dataset containing data fluctuation to perform fits to obtain parameter distribution, and we use data containing largely fluctuating points. The parameter values obtained by best fit with no largely fluctuating data are the most probable value. Therefore, the model parameters are identifiable. For other parameters, we use a 95% confidence interval of their non-linear fit or normal physical range. See Table S4 for a detailed summary of methods that give the uncertainty for each parameter and table S6-7 for best fit and uncertainties of parameters for all cases.

### Simulation of D-dimer dynamics with different initial T cell concentration

To study how different initial T cell values, *T*_0_, give different evolutions of D-dimer traces and thus influence the disease severity, we assume a linear dependence of T cell activation rate *δ*and inflammation inhibition rates, *λ, v*_*d*_, *v*_*h*_with *T*_0_. The parameters include T cell activation rate *δ*, and inflammation inhibition rates, *λ, v*_*d*_, *v*_*h*_. When *T*_0_ = 0.39(for non-survivors, determined from Figure 4b), Eq. (13)-(16) give non-survivors’ parameter values. When *T*_0_ = 0.84 (for survivors, determined from Figure 4b), Eq. (13)-(16) give survivors’ parameter values. Simulation in Figure 5c is performed using a series of *T*_0_ giving the corresponding *δ, λ, v*_*d*_, *v*_*h*_. Other parameters take fixed values of those of non-survivors.

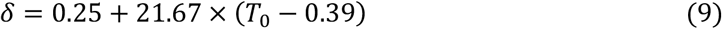

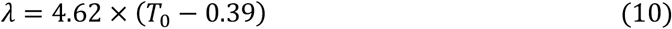

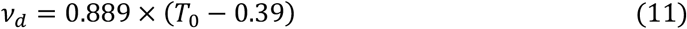

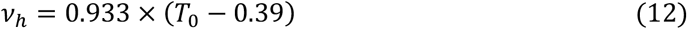

## Supporting information

A collection of supplementary figure, tables and texts

## Data Availability

All the codes to produce figures in our manuscript can be accessed using the following link

https://github.com/luhaozhang/covid19.git

## Acknowledgments

The research is funded by the W.M. Keck Foundation through award no. 1005586. The authors acknowledge Prof. Kelvin To for sharing data; Xiaoquan Wang, Guanghui She for discussion; Xinzi Zhang for writing suggestions.

