## Supplementary material for "Impairment of T cells’ antiviral and anti-inflammation immunities dominates death from COVID-19": A collection of supplementary figure, tables and texts

Luhao Zhang[1,2]†, Rong Li[1,3]†, Gang Song[4], Gregory D. Scholes[2]* and Zhen-Su She[1,3]*
[1]Institute of Health System Engineering, College of Engineering, Peking University, Beijing, China, 100871.
[2]Department of Chemistry, Princeton University, Princeton, NJ, 08540.
[3]State Key Laboratory for Turbulence and Complex Systems, Peking University, Beijing, China, 100871.
[4]Beijing Hospital, National Center of Gerontology; Institute of Geriatric Medicine, Chinese Academy of Medical Sciences, Beijing, China, 100730
†These authors contribute equally
*Corresponding author: Gregory D. Scholes, 609-258-0729,, 125, Frick Laboratory, Department of Chemistry, Princeton University, Princeton, NJ, and Zhen-Su She, 010-62766559,, 206, Peking University State Key Laboratory of Turbulence and Complex Systems, Beijing, China.

**This file includes:**

Supplementary text
Figures S1
Tables S1 to S12
SI References

### Supplementary Information Text

#### Asymptotic analysis of viral load peak

For viral dynamics Eq. (1)-(3), there are a total of six factors, the effective viral increasing rate $\tilde{\alpha}$, T cell clearance rate $\beta$, and death rate $\epsilon$, antibody preparation time $\tau$, antibody secretion rate $\eta$, and decay rate $\theta$. To obtain analytical solutions that how main factors determine viral peak, the decay of effector T cells and the production of antibodies are neglected, and Eq. (1)-(3) becomes,

$$\frac{dV(t)}{dt} = [\tilde{\alpha} - \beta T_{\mathrm{e}}(t)\,]V(t) \tag{S1}$$

$$\frac{dT_{\mathrm{e}}(t)}{dt} = \delta V(t) \tag{S2}$$

The analytical solutions are:

$$V(t) = V^* \frac{4}{2 + e^{-\tilde{\alpha}(t-t^*)} + e^{\tilde{\alpha}(t-t^*)}}, V^* = \frac{\tilde{\alpha}^2}{2\beta\delta} \tag{S3}$$

$$T_{\mathrm{e}}(t) = 2\tilde{\alpha}\frac{1}{1 + e^{-\tilde{\alpha}(t-t^*)}} \tag{S4}$$

where, $V^*$ is viral load peak described in the first section of Results, and $t^*$ is the corresponding time. The analytical results for all cases are summarized in Table S3 in SI, which shows they agree with numerical simulation with >90% overlap.

#### Parameter fixed during the fitting

For all the fits, $B_0$ is fixed to be cut-off values or to estimated values based on the antibody data points before the rise. $T_0$ is fixed at the maximum of data. $N$ is selected to be -5 or -10 according to different viral load profiles. $I_0$ for survivors is the value of the first three data points; $I_0$ for non-survivors is the mean value of the first three data points. For both survivors and non-survivors, $S_{\mathrm{d0}}$ was set to be 0.25 (half of the normal range), $S_{\mathrm{h0}}$ was set to be 0. $\epsilon$ and $\theta$ are the decay rates and were obtained by linear regression of the decay profiles. For cases without decay data points, the fitted $\epsilon$ values of other patients with the same severity were used, while an average of the fitted $\theta$ (P1, P3, 902, 916) 0.036 is used. For No.910 patient and non-survivors lacking saturated points for antibodies, only $\gamma$ and $\tau$ were varied. $\kappa$ for survivors is fixed equally to $\kappa$ of the best fit of non-survivors.

**Derivation of antiviral-inflammation equations for a group of individuals**

Taking viral load evolution as an example, the evolution of the mean of each variable over a group of individuals is written as:

$$\frac{dV_G}{dt} = \alpha_G V_G - \beta_G T_g V_G - \gamma_G A_G V_G + < \Delta\alpha_i \Delta V_i >$$
$$-\beta_G < \Delta T_i \Delta V_i > - < \Delta\beta_i \Delta T_i > V_G - < \Delta\beta_i \Delta V_i > T_G - < \Delta\beta_i \Delta T_i \Delta V_i >$$
$$-\gamma_G < \Delta A_i \Delta V_i > - < \Delta\gamma_i \Delta A_i > V_G - < \Delta\gamma_i \Delta V_i > A_G - < \Delta\gamma_i \Delta A_i \Delta V_i > \quad \text{(S5)}$$

Where, $V_G, T_g, A_G, \alpha_G, \beta_G, \gamma_G$ is the mean value of $V$, $T_e$, $A$, $\alpha, \beta, \gamma$ for a group of individuals. $\Delta V_i, \Delta T_i$, $\Delta A_i$, $\Delta\alpha_i$, $\Delta\beta_i$ and $\Delta\gamma_i$ is the deviations of the i$^{th}$ individual's variables and parameters. $< \Delta\alpha_i \Delta V_i >$ means the mean over all individual's $\Delta\alpha_i \Delta V_i$, so as other similar terms.

We assume, firstly, the deviation of variable and deviation of parameters are independent of each other $< \Delta\alpha_i \Delta V_i > \approx < \Delta\alpha_i >< \Delta V_i > = 0$, and $< \Delta\beta_i \Delta T_i > \approx 0, < \Delta\gamma_i \Delta V_i > \approx 0$. The deviation of an individual's $V_i$ changes across several scales over time and is the most prominent around $V_G$ maximum. Maximum of $\Delta V_i$, $\Delta T_i$ and $\Delta A_i$ are not overlap along time and $\Delta V_i, \Delta T_i, \Delta A_i$is not correlated; therefore, $< \Delta T_i \Delta V_i > \approx 0$, $< \Delta A_i \Delta V_i > \approx 0, < \Delta\alpha_i \Delta V_i > \approx 0$. We get:

$$\frac{dV_G}{dt} = \alpha_G V_G - \beta_G T_g V_G - \gamma_G A_G V_G \quad \text{(S6)}$$

In our fit for groups, we fit the median values of each variable of the group as an approximation to the mean values.

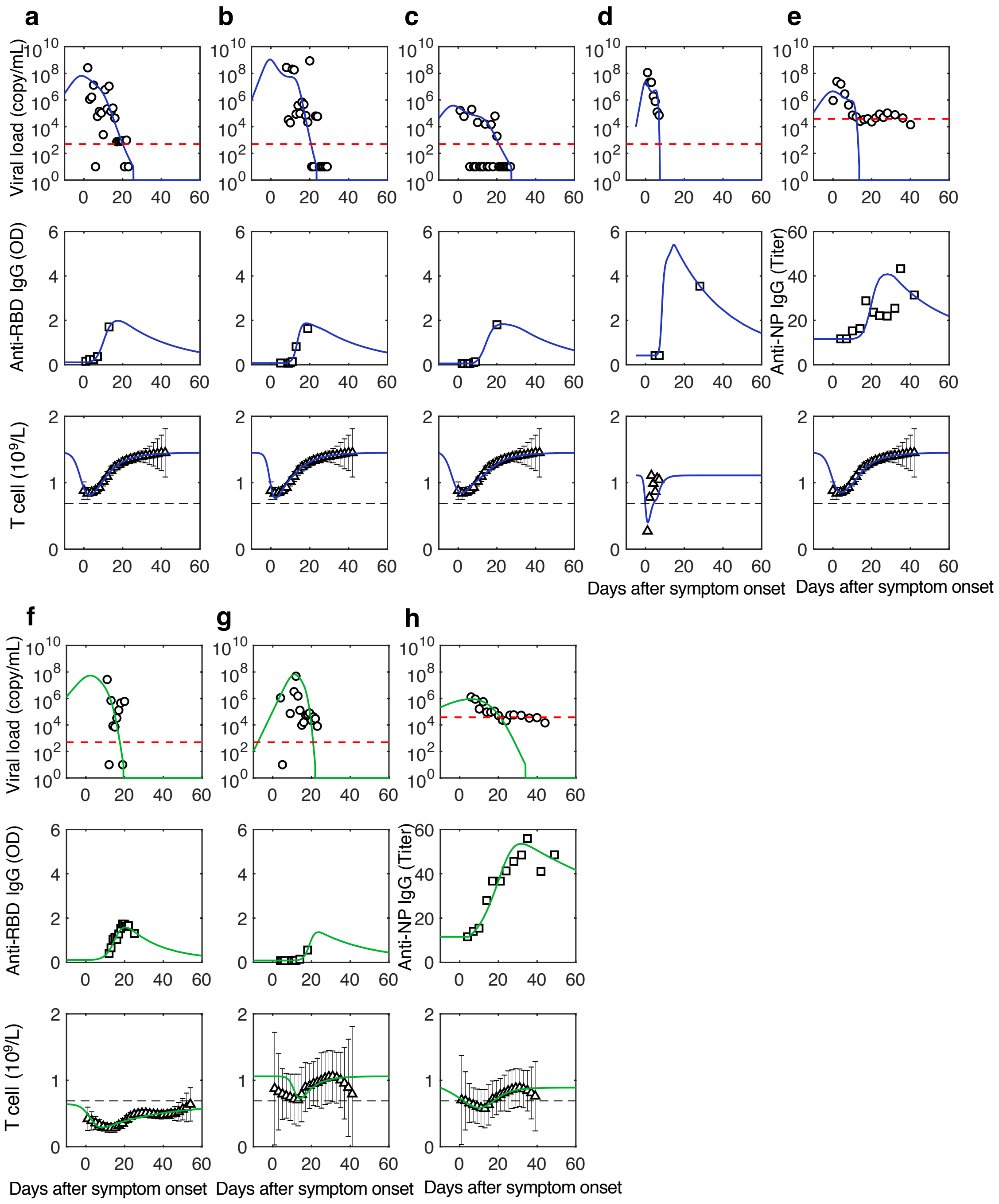

**Figure S1. Comparison of simulation of antiviral dynamics to data.** For dynamics of virus (circle), antibodies (square), and T cell count (triangle). a-e are mild individuals and the mild group, plotted in blue. f-h are severe(critical) individuals and the severe group, plotted in green.

**Table S1 Parameter names of the Antiviral-Inflammation Model and their units.**

| Parameter | Description | Unit |
|---|---|---|
| Antiviral parameters | | |
| $V_0$ | Initial viral load | copy/mL |
| $T_0$ | Initial naïve T cell concentration | $10^9$cell/L |
| $A_0$ | Initial antibody concentration | OD, or OD Ratio, or Titer |
| N | Starting day for the simulation | Days after symptom onset |
| $\tilde{\alpha}$ | Virus replication rate | $Day^{-1}$ |
| $\beta$ | Virus killing rate per unit T cells | $(10^9 cell/L)^{-1}$ $Day^{-1}$ |
| $\gamma$ | Virus killing rate by per unit antibodies | (OD, or OD Ratio, or Titer)$^{-1}$ $Day^{-1}$ |
| $\delta$ | T cell activation rate per unit virus | $(copy/mL)^{-1}(10^9 cell/L)$ $Day^{-1}$ |
| $\epsilon$ | T cell decay rate | $Day^{-1}$ |
| $\eta$ | Antibody secretion rate per unit virus | $(copy/mL)^{-1}$(OD, or OD Ratio, or Titer) $Day^{-1}$ |
| $\tau$ | Preparation time before antibody secretion begins | Day |
| $\theta$ | Antibody decay rate | $Day^{-1}$ |
| Inflammation parameters | | |
| $I_0$ | Initial concentration of IL-6 | pg/mL |
| $S_{d0}$ | Initial concentration of d-dimer | μg/mL |
| $S_{h0}$ | Initial concentration of high-sensitivity cardiac troponin I | ng/mL |
| $\kappa$ | IL-6 formation rate | (OD, or OD Ratio, or Titer)$^{-1}$ (pg/mL) $Day^{-1}$ |
| $\lambda$ | IL-6 inhibition rate | $Day^{-1}$ |
| $\mu_d$ | Formation rate of d-dimer | $(pg/mL)^{-1}(\mu g/mL)$ $Day^{-1}$ |
| $\nu_d$ | Inhibition rate of d-dimer | $Day^{-1}$ |
| $\mu_h$ | Formation rate of high-sensitivity cardiac troponin I | $(pg/mL)^{-1}(ng/mL)$ $Day^{-1}$ |
| $\nu_h$ | Inhibition rate of high-sensitivity cardiac troponin I | $Day^{-1}$ |

**Table S2 Populations of virus, T cell, and antibody data source integrated for mild, severe (critical), and non-survivor groups.**

| | Viral load | T cell | Antibody |
|---|---|---|---|
| 4 individuals from Isabella[28] | Isabella[28]'s cohort | Isabella[28]'s cohort (Lymphocyte count) | Isabella[28]'s cohort (Anti-S1 IgG) |
| 6 individuals from Kelvin[50] | Kelvin's cohort | Median values of CD3+ count of the mild group of HongzhouLu's cohort are used for 3 mild individuals, and median values of CD3+ count of the severe group of HongzhouLu's cohort are used for 3 severe individuals. | Kelvin's cohort |
| Group | | | |
| Mild | 38 individuals from Guohong Deng[1]' s nasopharyngeal swab data from the non-severe group. | 138 individuals from Hongzhou Lu[2]'s CD3+ count(mild group). | 22 individuals from Guohong Deng's Anti-NP IgG data for the non-severe group. |
| Severe (critical) | 20 severe + 9 critical individuals from Guohong Deng's nasopharyngeal swab data from the severe group. | 12 severe and 16 critical individuals from Hongzhou Lu's CD3+ count. Take the weighted average of severe and critical medians according to 20:9, so as the estimation of error bars. | 23 individuals from Guohong Deng's Anti-NP IgG from the severe group. |
| Survivors | Median data of 20 survivors from Kelvin To's cohort[50]. | 137 survived individuals from BinCao[3]'s Lymphocyte count. | Median data of 20 survivors from Kelvin To's cohort[50]. |
| Non-survivors | 2 individuals from Kelvin To[4]'s saliva data and Yazdan Yazdanpanah[5]'s nasopharyngeal data. | 54 individuals from BinCao[3]'s Lymphocyte count. | Use one of the 2 patients to represent the two with viral load data. |

**Table S3 Comparison of viral peak value obtained from Eq.S3 and simulation.**

Patient IDs for the patients described in Figure 2 and Figure S1 are provided. For example, a:908 means the patient described in Figure S1a is labelled as No.908; 2d: 902 means the patient described in Figure 2 is labelled as No.902).

| Patient ID | Eq.S3 (log10 scale) | Simulation (log10 scale) | value of Eq. S3/value of simulation (log10 scale) |
|---|---|---|---|
| **a:908** | 7.70 | 7.82 | 0.99 |
| **b:904** | 8.98 | 9.04 | 0.99 |
| **c:901** | 5.44 | 5.58 | 0.97 |
| **2d:902** | 6.74 | 6.80 | 0.99 |
| **f:910** | 7.74 | 7.82 | 0.99 |
| **g:916** | 7.68 | 7.73 | 0.99 |
| **2a:P1** | 7.14 | 7.18 | 0.99 |
| **2b:P3** | 9.13 | 9.16 | 1.00 |
| **d:P4** | 7.28 | 7.38 | 0.99 |
| **2c:P5** | 7.38 | 7.62 | 0.97 |
| **Survivors** | 5.88 | 6.11 | 0.96 |
| **Non-survivors** | 6.55 | 6.58 | 1.00 |
| **e: Mild** | 6.54 | 6.65 | 0.98 |
| **h: Severe group** | 5.59 | 5.96 | 0.94 |

**Table S4  Methods that give uncertainty for each parameter.**

| Parameter | Methods |
|---|---|
| Viral dynamics | |
| $V_0$ | We use a similar approach for uncertainty estimation as used by Marchingo, J. M. et al. Science. 346, 1123–1127 (2014) that uncertainty comes from the fluctuation of medical data. Therefore, large fluctuated data of viral load, T cell, and antibody, which are eliminated when performing best fit, are added to perform fits with data fluctuation. Within each data type, large fluctuated points are added one by one from early to later day, each time a fit is performed. Also, cases including fluctuated points in more than one data type are considered. For example, if a patient has two large fluctuated viral data points and three large fluctuated T cell data points, then the total number of fits with data fluctuation is six (2*3). The range for each parameter is given by the fitted parameters from all the fits with data fluctuation and the best fit, labelled as lower bound and upper bound. Then, we provide 95%CI of each parameter when 95% fits have the parameter in that regime. |
| $\tilde{\alpha}$ | |
| $\beta$ | |
| $\gamma$ | |
| $\delta$ | |
| $\eta$ | |
| $\tau$ | |
| N | N has no uncertainty because it is initially fixed. |
| T cell and antibody dynamics | |
| $\epsilon$ | The 95%CI of the fit of the decay profiles is used for 95%CI uncertainty. If the number of T cell data points for an individual's fit is two, the uncertainty of $\epsilon$ was estimated by that of the corresponding group who shares the same severity. 95%CI of No.916 patient's $\theta$ was used for cases whose uncertainty cannot be obtained by fitting. |
| $\theta$ | |
| $T_0$ | The 95%CI of the group data of the day with the highest median is used. Individual $T_0$'s uncertainty is estimated by that of the corresponding group, which has the same severity as the individual. |
| $A_0$ | $A_0$'s uncertainty is supposed to be determined by instrumental error and is not estimated from the limited number of data points in the initial stage (before rising). |
| Inflammation dynamics | |
| $\kappa$ | We use the same approach similar to viral dynamics. The way to consider group median data fluctuation is to make one of the group median data be substituted by the upper/lower limit value of its error bar, keeping others unchanged, and then a fit is performed. The process is repeated until each data point is substituted in one of the fits. Then the upper bound, lower bound, and 95%CI of each parameter is obtained using the same approach |
| $\lambda$ | |
| $\mu_d$ | |
| $\nu_d$ | |
| $\mu_h$ | |
| $\nu_h$ | |

| | |
|---|---|
| $I(0)$ | as in viral dynamics. The uncertainty of $\kappa$ of survivors is the same as non-survivors. |
| $S_d(0)$, $S_h(0)$ | The range of $S_{d0}$ was estimated by the normal range of D-dimer (Ref.2 in SI). The range of $S_{h0}$ is estimated between 0 and the average normal upper limit in men and women (Ref.6 in SI). 95%CI is obtained by calculating 95% regime of the range. |

**Table S5 Indices of data points of virus, T cells, and antibodies used for performing best fits of each case.**

Patient ID follows the Latin letter of their corresponding panel in Supplementary Fig 1 and Fig.2. For group data, best fits were performed using median values. "Total" means all the data were used. "-" means the data were not used. For fits of $I$, $S_d$, and $S_h$, all the data were used.

| Patient ID | $V$ data | $T$ data | $A$ data |
|---|---|---|---|
| **a:908** | [1,4,11,13,14,15,16:19,21,22] | Total | Total |
| **b:904** | [1,4,5,9:12] | Total | Total |
| **c:901** | [1,2,4,7,11,14,17,24] | Total | Total |
| **2d:902** | [1:7] | [1:16] | Total |
| **f:910** | [1,5,12,13] | [1:16] | Total |
| **g:916** | [1,3,5:6] | Total | Total |
| **2a:P1** | [1:4,5:7] | [2:5] | Total |
| **2b:P3** | [1:2,4:8] | [1,3,4,6,8,9] | Total |
| **d:P4** | [1:7] | [1,5,7] | Total |
| **2c:P5** | [4:8] | [1,3,5,6] | Total |
| **Survivors** | [1:6,8:10,12,14:16,17:19,21:23] | Total | [1:21,25] |
| **Non-survivors** | [1:8] | - | Total |
| **e:Mild group** | [1:7] | Total | [1:4,6,10,11] |
| **h:Severe group** | [1:2,4,6:8] | Total | [1:10,12] |

**Table S6 Best fit (with parameter uncertainty) to each patient (group).**

LB: Lower bound of the uncertainty. UB: Upper bound of the uncertainty. “-” means the parameter is fixed or the quality of data is not able to perform the estimation. Patient 2d:902 has no reasonable fitting other than best fit.

| Patient ID | $\tilde{\alpha}$ | $log10$ $(V_0)$ | $\beta$ | $\gamma$ | $\delta/10^{-8}$ | $\epsilon/10^{-1}$ | $\eta/10^{-8}$ | $\tau$ | $\theta/10^{-2}$ | $T_0$ | $B_0$ | N |
|---|---|---|---|---|---|---|---|---|---|---|---|---|
| **a:908** | 0.51 | 6.47 | 1.18 | 0.65 | 0.22 | 1.14 | 0.44 | 11.70 | 3.57 | 1.45 | 0.11 | -10 |
| LB | 0.48 | 5.45 | 1.12 | 0.50 | 0.22 | - | 0.44 | 11.70 | - | - | - | - |
| UB | 0.52 | 6.47 | 1.21 | 0.65 | 2.35 | - | 4.87 | 12.36 | - | - | - | - |
| 95% CI | (LB, UB) | | | | | (1.06, 1.22) | (LB, UB) | | (0, 10.99) | (1.09, 1.81) | | |
| **b:904** | 0.89 | 5.98 | 2.08 | 1.21 | 0.02 | 1.14 | 0.04 | 14.09 | 3.57 | 1.45 | 0.08 | -10 |
| LB | 0.82 | 4.31 | 1.93 | 0.88 | 0.02 | - | 0.04 | 13.90 | - | - | - | - |
| UB | 1.09 | 5.98 | 2.50 | 1.21 | 0.37 | - | 0.65 | 14.56 | - | - | - | - |
| 95% CI | (LB, UB) | | | | | (1.06, 1.22) | (LB, UB) | | (0, 10.99) | (1.09, 1.81) | | |
| **c:901** | 0.47 | 4.62 | 1.15 | 0.54 | 34.92 | 1.14 | 68.40 | 17.18 | 3.57 | 1.45 | 0.06 | -10 |
| LB | 0.47 | 4.24 | 1.15 | 0.54 | 30.48 | - | 64.64 | 17.18 | 0 | - | - | - |
| UB | 0.59 | 4.62 | 1.44 | 0.64 | 34.92 | - | 68.40 | 18.43 | 10.99 | - | - | - |
| 95% CI | (LB, UB) | | | | | (1.06, 1.22) | (LB, UB) | | (0, 10.99) | (1.09, 1.81) | | |
| **2d:902** | 0.58 | 3.69 | 1.70 | 1.19 | 1.81 | 1.40 | 4.48 | 5.40 | 5.11 | 1.06 | 0.11 | -10 |
| LB | - | - | - | - | - | - | - | - | - | - | - | - |
| UB | - | - | - | - | - | - | - | - | - | - | - | - |
| 95% CI | (LB, UB) | | | | | (1.17, 1.64) | (LB, UB) | | (0, 10.99) | (0.68, 1.44) | - | - |

| | | | | | | | | | | | | |
|---|---|---|---|---|---|---|---|---|---|---|---|---|
| **f:910** | 0.70 | 2.00 | 3.00 | 3.36 | 0.15 | 1.40 | 0.40 | 7.74 | 3.57 | 1.06 | 0.08 | -10 |
| LB | - | - | - | 2.52 | - | - | 0.40 | 7.74 | - | - | - | - |
| UB | - | - | - | 4.68 | - | - | 2.08 | 10.80 | - | - | - | - |
| 95% CI | (LB, UB) | | | | | (1.17, 1.64) | (LB, UB) | | (0, 10.99) | (0.68, 1.44) | - | - |
| **g:916** | 0.41 | 6.12 | 1.93 | 1.44 | 0.09 | 0.33 | 0.43 | 12.79 | 5.49 | 0.64 | 0.11 | -10 |
| LB | 0.33 | 5.27 | 1.50 | 0.10 | 0.06 | - | 0.28 | 12.79 | - | - | - | - |
| UB | 0.46 | 6.66 | 2.28 | 1.75 | 0.65 | - | 3.28 | 13.83 | - | - | - | - |
| 95% CI | (LB, UB) | | | | | (0.22, 0.44) | (LB, UB) | | (0, 10.99) | (0.39, 0.89) | - | - |
| | $\tilde{\alpha}$ | $log10(V_0)$ | $\beta$ | $\gamma$ | $\delta/10^{-8}$ | $\epsilon/10^{-1}$ | $\eta/10^{-8}$ | $\tau$ | $\theta/10^{-2}$ | $T_0$ | $B_0$ | N |
| **2a:P1** | 1.17 | 2.93 | 10.00 | 1.52 | 0.50 | 1.14 | 14.58 | 11.22 | 1.65 | 1.74 | 0.30 | -5 |
| LB | 0.79 | 2.93 | 7.18 | 1.52 | 0.07 | - | 1.91 | 11.02 | - | - | - | - |
| UB | 1.17 | 7.70 | 10.00 | 2.58 | 0.57 | - | 14.58 | 18.62 | - | - | - | - |
| 95% CI | (LB, UB) | | | | | (1.06, 1.22) | (LB, UB) | | (0, 10.99) | (1.53, 1.95) | - | - |
| **2b:P3** | 1.48 | 6.63 | 8.05 | 4.72 | 0.01 | 0.78 | 0.02 | 20.13 | 2.03 | 0.90 | 0.16 | -10 |
| LB | 1.30 | 6.61 | 8.05 | 4.72 | 0.01 | - | 0.02 | 20.10 | - | - | - | - |
| UB | 1.48 | 7.01 | 8.82 | 4.91 | 0.01 | - | 0.03 | 20.48 | - | - | - | - |
| 95% CI | (LB, UB) | | | | | (0.76, 0.80) | (LB, UB) | | (0, 10.99) | (0.69, 1.11) | - | - |
| **d:P4** | 1.86 | 4.00 | 3.84 | 47.30 | 2.38 | 4.14 | 7.84 | 8.45 | 3.57 | 1.11 | 0.42 | -5 |
| LB | 1.83 | 3.69 | 3.84 | 43.57 | 1.65 | - | 7.56 | 8.45 | - | - | - | - |
| UB | 2.11 | 4.29 | 7.02 | 49.25 | 2.38 | - | 8.83 | 8.83 | - | - | - | - |
| 95% CI | (LB, UB) | | | | | (4.11, | (LB, UB) | | (0, | (0.90, | - | - |

| | | | | | | | | | | | | |
|---|---|---|---|---|---|---|---|---|---|---|---|---|
| | | | | | | 5.01) | | | 10.99) | 1.32) | | |
| **2c:P5** | 0.82 | 4.69 | 1.31 | 0.11 | 1.08 | 4.72 | 9.26 | 6.55 | 3.57 | 0.88 | 0.10 | -5 |
| LB | 0.70 | 4.69 | 1.14 | 0.11 | 0.91 | - | 8.12 | 6.55 | - | - | - | - |
| UB | 1.13 | 5.74 | 1.75 | 0.14 | 1.60 | - | 12.86 | 9.60 | - | - | - | - |
| 95% CI | (LB, UB) | | | | | (3.43, 4.80) | (LB, UB) | | (0, 10.99) | (0.66, 1.11) | - | - |
| **Survivor Group** | 0.67 | 5.34 | 2.96 | 1.16 | 10.00 | 3.21 | 18.37 | 9.70 | 3.51 | 0.84 | 0.13 | 0 |
| LB | 0.67 | 5.20 | 2.96 | 1.11 | 9.50 | 2.68 | 17.22 | 9.28 | - | - | - | - |
| UB | 0.79 | 5.34 | 3.59 | 1.24 | 10.00 | 3.21 | 19.59 | 9.85 | - | - | - | - |
| 95% CI | (LB, UB) | | | | | (2.68, 3.21) | (LB, UB) | | (0, 10.99) | (0.72, 0.92) | - | - |
| **Non-survivor Group** | 0.55 | 5.00 | 16.98 | 3.28 | 0.25 | 0.33 | 23.00 | 7.98 | 0.00 | 0.39 | 0.08 | 0 |
| LB | - | - | - | - | - | 0.22 | - | - | - | - | - | - |
| UB | - | - | - | - | - | 0.44 | - | - | - | - | - | - |
| 95% CI | | | | | | (0.22, 0.44) | - | - | - | (0.29, 0.49) | - | - |
| | $\tilde{\alpha}$ | $log10(V_0)$ | $\beta$ | $\gamma$ | $\delta/10^{-8}$ | $\epsilon/10^{-1}$ | $\eta/10^{-7}$ | $\tau$ | $\theta/10^{-2}$ | $T_0$ | $B_0$ | N |
| **e: Mild Group** | 0.51 | 5.07 | 1.25 | 7.83 | 0.30 | 1.14 | 9.81 | 20.00 | 3.57 | 1.45 | 11.67 | -10 |
| LB | 0.41 | 4.82 | 1.00 | 2.58 | 2.81 | - | 6.98 | 19.30 | - | - | - | - |
| UB | 0.60 | 5.76 | 1.44 | 7.83 | 4.16 | - | 9.81 | 20.00 | - | - | - | - |
| 95% CI | (LB, UB) | | | | | (1.06, 1.22) | (LB, UB) | | (0, 10.99) | (1.09, 1.81) | | |

| | | | | | | | | | | | | |
|---|---|---|---|---|---|---|---|---|---|---|---|---|
| **h: Severe Group** | 0.16 | 5.32 | 0.52 | 0.02 | 0.63 | 1.56 | 30.93 | 14.61 | 1.29 | 0.89 | 11.53 | -10 |
| LB | 0.15 | 2.63 | 0.50 | 0.02 | 5.93 | - | 30.60 | 13.58 | - | - | - | - |
| UB | 0.61 | 5.32 | 2.51 | 0.16 | 8.49 | - | 39.99 | 15.05 | - | - | - | - |
| 95% CI | (LB, UB) | | | | | (1.28, 1.84) | (LB, UB) | | (0, 10.99) | (0.60, 1.17) | - | - |

**Table S7 Best fit (with parameter uncertainty) for survivors and non-survivors.**

Uncertainty of $S_d(0)$ was estimated by the normal range of d-dimer[2]. The range of $S_h(0)$ is estimated between 0 and the upper limit of the first data point of HSCT. The upper reference limit of HSCT in Fig.4 was from the averaged values in men and women[6]. $I_0$ = 5.4, which is set to be the middle of the normal range of IL-6[7].

| **Survivors** | $I(0)$ | $S_{d0}(0)$ | $S_{h0}(0)$ | $\kappa$ | $\lambda$ | $\mu_d$ | $\nu_d$ | $\mu_h$ | $\nu_h$ |
|---|---|---|---|---|---|---|---|---|---|
| Best fit | 5.58 | 0.25 | 0 | 1.63 | 2.08 | 0.04 | 0.40 | 0.25 | 0.42 |
| LB | 3.60 | 0.00 | 0.00 | 0.11 | 1.20 | 0.04 | 0.40 | 0.24 | 0.40 |
| UB | 11.44 | 0.5 | 2.5 | 2.12 | 3.00 | 0.07 | 0.40 | 0.26 | 0.44 |
| 95%CI | (LB, UB) | (0.01, 0.49) | (0, 90.49) | (LB, UB) | | | | | |
| **Non-survivors** | $I(0)$ | $S_{d0}(0)$ | $S_{h0}(0)$ | $\kappa$ | $\lambda$ | $\mu_d$ | $\nu_d$ | $\mu_h$ | $\nu_h$ |
| Best fit | 10.73 | 0.25 | 0.00 | 1.63 | 0.00 | 0.08 | 0.00 | 0.37 | 0.00 |
| LB | 9.25 | 0.00 | 0.00 | 0.11 | 0.00 | 0.06 | 0.00 | 0.32 | 0.00 |
| UB | 12.83 | 0.50 | 95.25 | 2.12 | 0.02 | 0.11 | 0.00 | 1.00 | 0.37 |
| 95%CI | (LB, UB) | (0.01, 0.49) | (0, 90.49) | (LB, UB) | | | | | |

**Table S8 Equation for goodness of fit (F) for each variable.**

$V_{\mathrm{max}}, T_{\mathrm{max}}, A_{\mathrm{max}}, L_{\mathrm{max}}$ are the maximums of the simulations for the virus, T cells, antibodies, and inflammation markers. $n_V, n_T, n_L$ are the number of data points. $V_i, T_i, A_i, L_i$ are the value of data and $\bar{V}_i, \bar{T}_i, \bar{A}_i, \bar{L}_i$ are the simulated value.

| Variable | Equations of $F$ |
|---|---|
| ***V*** | $1-\frac{1}{\log 10(V_{\mathrm{max}})}\sqrt{\frac{1}{n_V}\sum_{i=1}^{n_V}(\log10 V_i-\log10 \bar{V}_i)^2}$ |
| ***T*** | $1-\frac{1}{T_{\mathrm{max}}}\sqrt{\frac{1}{n_T}\sum_{i=1}^{n_T}(T_i-\bar{T}_i)^2}$ |
| ***A*** | $1-\frac{1}{A_{\mathrm{max}}}\sqrt{\frac{1}{n_A}\sum_{i=1}^{n_A}(A_i-\bar{A}_i)^2}$ |
| ***I*, *S*d, *S*h for survivors** | $1-\frac{1}{L_{\mathrm{max}}}\sqrt{\frac{1}{n_L}\sum_{i=1}^{n_L}(L_i-\bar{L}_i)^2}\,, L=I, S_{\mathrm{d}}, S_{\mathrm{h}}$ |
| ***I*, *S*d, *S*h for non-survivors** | $1-\frac{1}{\log 10(L_{\mathrm{max}})}\sqrt{\frac{1}{n_L}\sum_{i=1}^{n_L}(\log10 L_i-\log10 \bar{L}_i)^2}\,, L=I, S_{\mathrm{d}}, S_{\mathrm{h}}$ |

**Table S9 Summary of the goodness of fit (F)**

| | *V* | *T* | *A* | *I* | $S_d$ | $S_h$ |
|---|---|---|---|---|---|---|
| **a:908** | 0.92 | 0.99 | 0.97 | - | | |
| **b:904** | 0.95 | 0.98 | 0.94 | | | |
| **c:901** | 0.94 | 0.98 | 0.99 | | | |
| **2d:902** | 0.85 | 0.94 | 0.89 | | | |
| **f:910** | 0.97 | 0.86 | 0.99 | | | |
| **g:916** | 0.91 | 0.94 | 0.92 | | | |
| **2a:P1** | 0.96 | 0.99 | 0.99 | | | |
| **2b:P3** | 0.97 | 0.94 | 0.96 | | | |
| **d:P4** | 0.89 | 0.86 | 0.97 | | | |
| **2c:P5** | 0.82 | 0.91 | 0.84 | | | |
| **Survivors** | 0.81 | 0.97 | 0.85 | 0.89 | 0.74 | 0.79 |
| **Non-survivors** | 0.77 | 0.84 | 0.98 | 0.97 | 0.97 | 0.95 |
| **e:Mild** | 0.90 | 0.97 | 0.89 | - | | |
| **h:Severe group** | 0.97 | 0.96 | 0.92 | | | |

**Table S10 Constraints of input parameters for each case.**

<table>
<tr><th>Patient ID</th><th>$\tilde{\alpha}$</th><th>$log10(V_0)$</th><th>$\beta$</th><th>$\gamma$</th><th>$\delta/10^{-8}$</th><th>$\epsilon/10^{-1}$</th><th>$\eta/10^{-8}$</th><th>$\tau$</th><th>$\theta/10^{-2}$</th></tr>
<tr><td>**a:908**</td><td rowspan="5">0.1<br>2.5</td><td rowspan="5">1<br>8</td><td rowspan="3">1<br>5</td><td rowspan="6">0.1<br>5</td><td rowspan="2">0.01<br>5</td><td rowspan="3">1.14</td><td rowspan="2">0.02<br>5</td><td rowspan="3">5<br>20</td><td rowspan="3">3.57</td></tr>
<tr><td>**b:904**</td></tr>
<tr><td>**c:901**</td><td>0.01<br>50</td><td>0.02<br>100</td></tr>
<tr><td>**2d:902**</td><td rowspan="2">1<br>10</td><td rowspan="2">0.01<br>5</td><td>1.40</td><td rowspan="2">0.02<br>5</td><td rowspan="3">1<br>20</td><td>5.11</td></tr>
<tr><td>**f:910**</td><td>0.33</td><td>5.49</td></tr>
<tr><td>**g:916**</td><td>0.7</td><td>2</td><td>3</td><td>0.15</td><td>1.40</td><td>0.4<br>5</td><td>3.57</td></tr>
<tr><td>**2a:P1**</td><td rowspan="3">0.1<br>2.5</td><td rowspan="5">1<br>8</td><td rowspan="3">1<br>10</td><td rowspan="2">0.1<br>5</td><td rowspan="3">0.01<br>5</td><td>1.14</td><td rowspan="3">0.02<br>20</td><td rowspan="3">5<br>25</td><td>1.65</td></tr>
<tr><td>**2b:P3**</td><td>0.30<br>0.90</td><td>2.03</td></tr>
<tr><td>**d:P4**</td><td>0.1<br>50</td><td>4.10<br>10.50</td><td>3.57</td></tr>
<tr><td>**2c:P5**</td><td>0.5<br>1.5</td><td>0.5<br>2</td><td>0.05<br>0.15</td><td>0.1<br>2.5</td><td>1<br>8</td><td>1<br>16</td><td>1<br>10</td><td>3.57</td></tr>
<tr><td>**Survivors**</td><td>0.1<br>2.5</td><td>1<br>5</td><td>0.1<br>5</td><td>1<br>10</td><td>1<br>5</td><td>0.02<br>20</td><td>5<br>20</td><td>3.51</td></tr>
</table>

| | | | | | | | | | |
|---|---|---|---|---|---|---|---|---|---|
| **Non-survivors** | 0.55 | 5 | 16.98 | 0.5<br>8 | 0.25 | 0.33 | 23 | 1<br>15 | 0.00 |
| | $\tilde{\alpha}$ | $log10(V_0)$ | $\beta$ | $\gamma$ | $\delta/10^{-8}$ | $\epsilon/10^{-1}$ | $\eta/10^{-7}$ | $\tau$ | $\theta/10^{-2}$ |
| **e: Mild Group** | 0.1<br>2.5 | 1<br>8 | 1<br>5 | 1<br>10 | 0.1<br>5 | 1.14 | 1<br>10 | 5<br>20 | 3.57 |
| **h: Severe Group** | | | 0.5<br>5 | 0.002<br>1 | 1<br>10 | 1.56 | 10<br>40 | | 1.29 |

**Table S11 Initial guess of parameters for each case.**

| Patient ID | $\tilde{\alpha}$ | $log10(V_0)$ | $\beta$ | $\gamma$ | $\delta$ $/10^{-8}$ | $\epsilon$ $/10^{-1}$ | $\eta$ $/10^{-8}$ | $\tau$ | $\theta$ $/10^{-2}$ | $T_0$ | $A_0$ | N |
|---|---|---|---|---|---|---|---|---|---|---|---|---|
| **a:908** | 0.80 | 6.00 | 2.00 | 0.80 | 0.07 | 1.14 | 0.16 | 8.00 | 3.57 | 1.45 | 0.11 | -10 |
| **b:904** | 0.80 | 6.00 | 1.60 | 1.00 | 0.02 | 1.14 | 0.03 | 15.00 | 3.57 | 1.45 | 0.08 | -10 |
| **c:901** | 0.50 | 4.00 | 1.40 | 0.50 | 30.00 | 1.14 | 70.00 | 15.00 | 3.57 | 1.45 | 0.06 | -10 |
| **2d:902** | 1.00 | 1.50 | 4.50 | 1.12 | 0.50 | 1.40 | 1.80 | 4.50 | 5.11 | 1.06 | 0.11 | -10 |
| **f:910** | 1.00 | 2.00 | 6.00 | 2.40 | 0.18 | 0.33 | 0.95 | 6.00 | 5.49 | 0.64 | 0.11 | -10 |
| **g:916** | 0.70 | 2.00 | 3.00 | 2.50 | 0.15 | 1.40 | 0.50 | 7.50 | 3.57 | 1.06 | 0.08 | -10 |
| **2a:P1** | 1.00 | 4.00 | 4.92 | 0.30 | 0.59 | 1.14 | 11.00 | 10.00 | 1.65 | 1.74 | 0.30 | -5 |
| **2b:P3** | 1.30 | 7.00 | 4.58 | 2.00 | 0.01 | 0.60 | 0.03 | 18.00 | 2.03 | 0.90 | 0.16 | -5 |
| **d:P4** | 1.60 | 5.00 | 3.06 | 1.20 | 0.35 | 4.10 | 1.20 | 14.00 | 3.57 | 1.11 | 0.42 | -5 |
| **2c:P5** | 0.80 | 4.00 | 1.10 | 0.09 | 0.29 | 4.70 | 4.00 | 3.50 | 3.57 | 0.88 | 0.10 | -5 |
| **Survivor-s** | 1.00 | 5.00 | 2.60 | 0.90 | 4.00 | 0.15 | 9.00 | 9.50 | 3.57 | 0.84 | 0.13 | 0 |
| **Non-survivors** | 0.55 | 5.00 | 16.98 | 4.00 | 0.25 | 0.33 | 23.00 | 8.00 | 0.00 | 0.39 | 0.08 | 0 |
| | $\tilde{\alpha}$ | $log10(V_0)$ | $\beta$ | $\gamma$ | $\delta$ $/10^{-8}$ | $\epsilon$ $/10^{-1}$ | $\eta$ $/10^{-7}$ | $\tau$ | $\theta$ $/10^{-2}$ | $T_0$ | $A_0$ | N |
| **e:Mild Group** | 0.86 | 4.00 | 2.51 | 1.00 | 1.00 | 1.14 | 1.00 | 10.00 | 4.57 | 1.45 | 11.67 | -10 |
| **h:Severe Group** | 0.3 | 5.8 | 1.20 | 0.01 | 6 | 1.56 | 27 | 10.00 | 1.29 | 0.89 | 11.53 | -10 |

**Table S12 Constraints of parameters to be varied and initial guesses for best fits of survivors and non-survivors.**

"-" means the parameter is fixed during the fit.

| | $I(0)$ | $S_d(0)$ | $S_h(0)$ | $\kappa$ | $\lambda$ | $\mu_d$ | $\nu_d$ | $\mu_h$ | $\nu_h$ |
|---|---|---|---|---|---|---|---|---|---|
| **Survivors** | - | - | - | 1.63<br>1.63 | 0<br>3 | 0<br>1 | 0.4<br>0.6 | 0<br>1 | 0.4<br>0.6 |
| Initial guess | 6.40 | 0.25 | 0 | 1.63 | 2.32 | 0.04 | 0.40 | 0.25 | 0.42 |
| | $I(0)$ | $S_d(0)$ | $S_h(0)$ | $\kappa$ | $\lambda$ | $\mu_d$ | $\nu_d$ | $\mu_h$ | $\nu_h$ |
| **Non-survivors** | - | - | - | 0<br>3 | 0<br>3 | 0<br>1 | 0<br>1 | 0<br>1 | 0<br>1 |
| Initial guess | 11.73 | 0.25 | 0 | 1.82 | 0.00 | 0.085 | 0.00 | 0.38 | 0.00 |
